## Supplementary material for "Evaluating COVID-19 booster vaccination strategies in a partially vaccinated population: a modeling study"


**Supplementary material** **1**
Transmission model 1
Overview of the model 1
Modeling of vaccination 2
Statistical framework 3
Estimation of hospital-related outcomes 4
Supplementary results 6
Primary vaccination vs booster doses 6
Sensitivity analysis with 100,000 vaccine doses administered daily 8
Sensitivity analysis with 400,000 vaccine doses administered daily 11
Model fitting 14
Epidemiology 15
References 17


#### Transmission model

##### Overview of the model

We developed a deterministic, compartmental, ordinary differential equation (ODE) model, which has been enhanced in successive stages and described elsewhere.[1–3] It was age-structured to account for specific contact patterns between different age groups, using the inter-individual contact matrices estimated by Prem et al.[4] We considered 17 age groups: 16 groups of five-year span, and a final group for people aged 80 years and more. The population structure of each region was retrieved from 2017 census data provided by the French National Institute of Statistics and Economic Studies (Insee).[5]

Supplementary Figure S1 depicts the transmission model. We considered that individuals were susceptible (S), and then potentially exposed to the virus but not infectious (E). As observed in clinical practice, we set children and adolescents to be less susceptible to infection than other age groups.[6] Exposed individuals stay in their compartment for an average of 2.72 days[7] before moving to either asymptomatic (A) or pre-symptomatic (Ips) compartment according to observed risk of being asymptomatic.[6] We considered that asymptomatic individuals were 45% less infectious than pre-symptomatic individuals were[8] and we assumed they stayed in the compartment for an average of 10.91 days before moving to the removed compartment (R) of patients that are cured or dead from COVID-19. Pre-symptomatic individuals will become infected symptomatic (Is) after an average duration of 2.38 days. This choice of parameters gave a mean incubation period of 5.1 days,[9] within around 2 days of pre-symptomatic transmissions. Then they become after an average of 5 days (assumption) either hospitalized (Ih) or remain within the community (Inh) according to age-dependent hospitalization risks[10,11] adjusted to apply only to clinical cases.[6] We assumed that hospitalized individuals were no longer infectious because of their hospitalization whereas non-hospitalized keep spreading the disease with the same intensity as symptomatic individuals. To account for the impact of VOC, we considered a 40%-increase in the risk of hospitalization between Alpha VOC and wild type[12] and a 80%-increase between Delta VOC and Alpha VOC.[13] These modifications in the parameters were applied gradually starting one month prior to the date on which each VOC became dominant in the country (i.e. February 28th 2021 for Alpha VOC, and July 15th 2021 for Delta VOC). We assumed the average duration in the (Inh) compartment was 3.53 days to match the total duration in the asymptomatic compartment. Finally, both hospitalized and non-hospitalized individuals moved to the removed (R) compartment. Therefore, we assume no waning of immunity.

To match the epidemic dynamics and reproduce the Erlang distributions of durations in each compartment of the transmission model, we subdivided each compartment having a role in the infection process into 10 sub-compartments. This subdivision had no impact on the mean duration spent in each compartment.

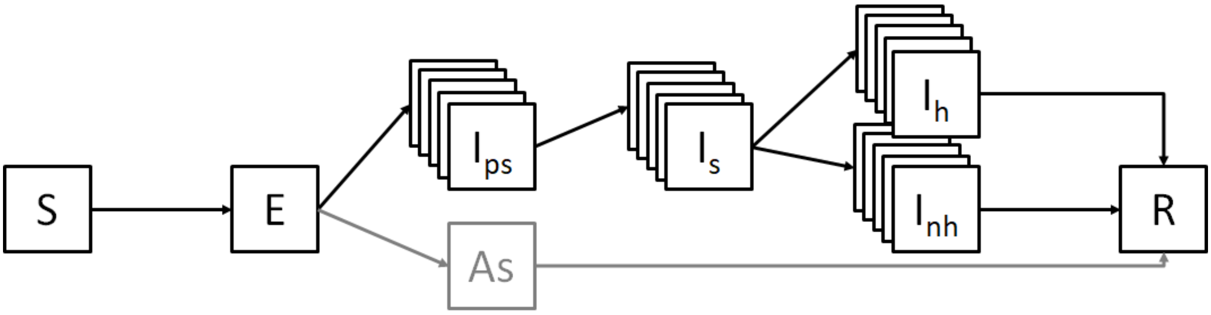


**Supplementary Figure S1** Diagram of the SARS-CoV-2 transmission model. **S:** susceptible, **E:** exposed, **Ips**: infectious pre-symptomatic, **Is:** infectious symptomatic, **Ih**: infectious hospitalized, **Inh**: infectious non-hospitalized, **As**: asymptomatic, **R**: removed.

**Modeling of vaccination**

To include the process of vaccination, all compartments and transition flows are duplicated into parallel branches that represent the mRNA or vector vaccines (Supplementary Figure S2). We assumed only individuals in the $S$ or $R$ compartment could be vaccinated. Upon vaccination with a mRNA vaccine (or "vector" vaccine), individuals moved from their current compartment in the main branch to its counterpart on the parallel "mRNA" branch (or "vector" branch). Once in a vaccine-specific branch, they were subject to the same transitions as in the main branch, but with different parameters modified according to the vaccines' efficacies on infection, symptomatic cases and hospitalization.

We designed a framework to translate real-life vaccination strategies into parameters that can be used in our compartmental model. The vaccination strategies were first constructed in terms of proportion of population vaccinated at some time points by age group (i.e. vaccination coverage targets). These proportions were then transcribed into rates to be used into the transmission model. The framework made it possible to compute rates from any combination of vaccination strategies, any number of coverage goals, and any number of population groups. This allowed us to first, reproduce the actual vaccination campaign as it happened in France, and second, to model the different vaccination strategies to assess from September 1st, 2021 onwards.

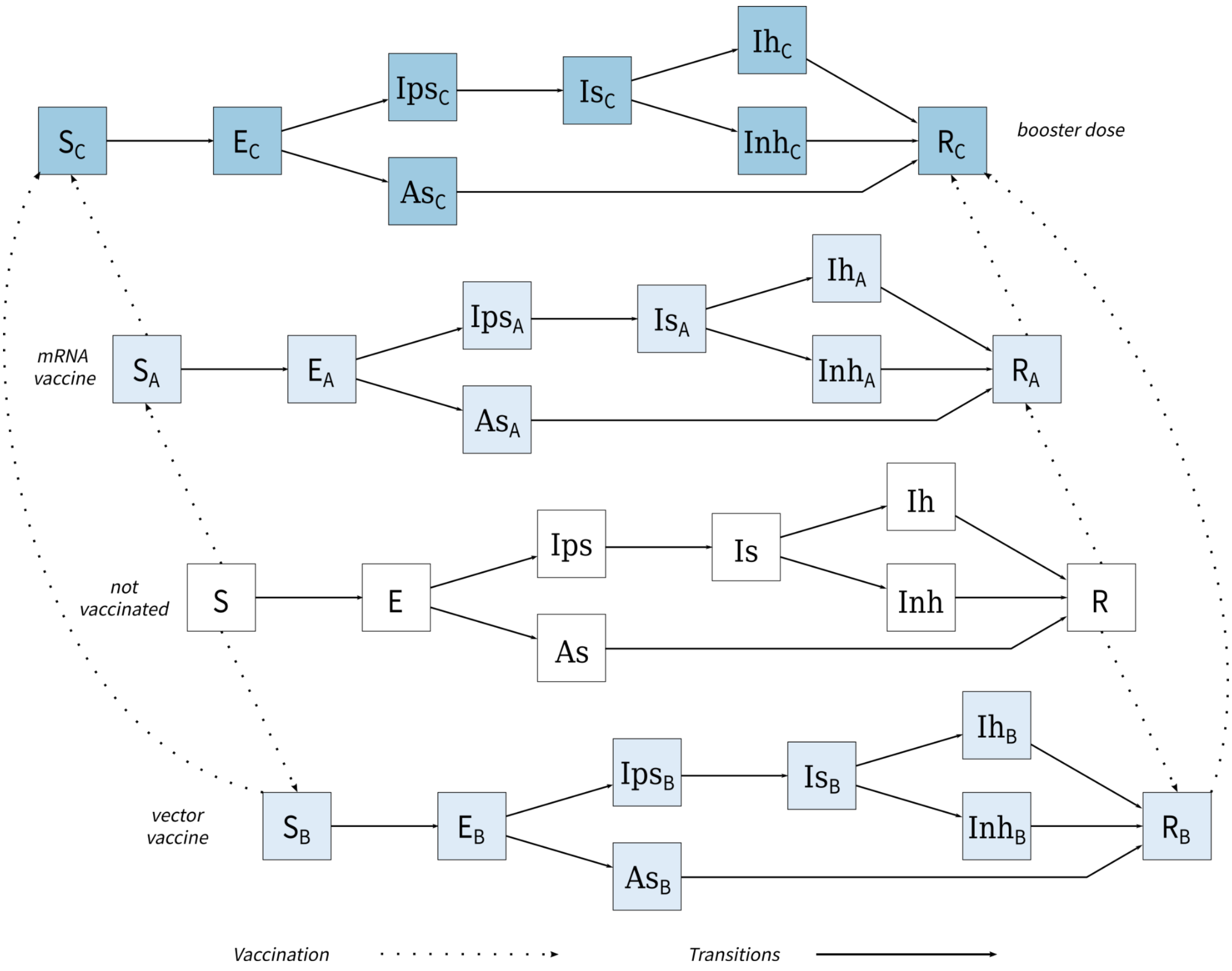


**Supplementary Figure S2** Diagram of the multi-branch SARS-CoV-2 transmission model taking into account vaccines.

**Statistical framework**

We retrieved publicly available epidemiological regional data related to the COVID-19 epidemic in metropolitan France gathered by the French National Public Health Agency (SpF-'Santé publique France')[14] daily number of hospital admissions (general and intensive care unit (ICU) wards), daily number of ICU admissions, daily number of occupied ICU beds, and daily number of deaths in hospitals (deaths in nursing homes and at home were not considered). These epidemiological data were corrected for reporting delays following the same procedure as Salje et al.[15]

Based on these hospitalization data, we estimated model parameters by maximum likelihood in a three-step process using the *bbmle* R package.[16] First, on the period stretching from March 14 to May 10, 2020, corresponding to the evolution of the epidemic until the end of the first french national lockdown, we estimated the value of the transmission parameter β, governing the value of the basic reproduction number R0, the initial state in each compartment per age group on March 1, 2020 and the effects of the national lockdown. The latter was estimated through a transmission reduction parameter, hereafter called $c_\beta$, and multiplied the transmission parameter (and thus contact matrices) to reproduce the several mitigation measures implemented, and their consequences on the regional propagation. We jointly estimated these parameters by fitting the daily deseasonalized time series of hospital admissions (hereafter denoted $Hosp$) using the likelihood $L_\beta$ defined in Equation (5.1).

$$L_{\beta} = \prod_{t} NBin\left(Hosp_{observed}(t) | Hosp_{predicted}(t)\right) \qquad (5.1)$$

where $NBin(.|X)$ is a negative binomial distribution of mean $X$ and overdispersion $X^{\delta}$, δ being a parameter specific to each region to be estimated. Confidence intervals of these two parameters were estimated using likelihood profiling methods.

In a second step, we jointly estimated three regional coefficients adjusting age-specific risks of ICU admissions, risk of deaths and lengths of stay in ICU (including ICU stays leading to death). They were estimated by simultaneously fitting the time series of ICU admissions (hereafter denoted ICU), deaths (both smoothed using 7-day centered moving average) and occupied ICU beds (hereafter denoted BedICU) from March 14 to May 10, 2020 using the likelihood defined in Equation (5.2).

$$L_{ICU-deaths} = \prod_{t} NBin\left(ICU_{observed}(t) | ICU_{predicted}(t)\right) \cdot NBin\left(deaths_{observed}(t) | deaths_{predicted}(t)\right) \cdot NBin\left(BedICU_{observed}(t) | BedICU_{predicted}(t)\right) \qquad (5.2)$$

where $NBin(.|X)$ is a negative binomial distribution of mean $X$ and overdispersion $X^{\delta}$, δ being a parameter specific to each region to be estimated.

Finally, starting from May 11, 2020, we estimated the transmission reduction parameter on 7-day moving time windows. If we note *J* the day of data update,  was estimated by fitting the daily deseasonalized time series of hospital admissions from *J-6* till *J* included, while it corresponded to mitigation measures implemented over the period stretching from *J-13* to *J* included (14 days in total). We renewed the calibration every 3 days, which means that at each new calibration the preceding calibration was kept from *J-13* and *J-11* and partly replaced from *J-10* to *J* by the newly estimated value.

The model itself was implemented in the *C++* programming language[17] and the analysis was made using the *R* programming language.[18]

**Estimation of hospital-related outcomes**

Based on the estimated number of new hospital admissions per day provided by our epidemiological model, we inferred outcomes related to hospital requirements, namely ICU admissions, and deaths.

We divided the hospital settings in two parts: general ward and ICU ward (Supplementary Figure S3). The epidemiological transmission model estimated the daily number of new hospitalized cases due to COVID-19 infection, regardless of the ward (i.e. ICU and general wards). Once admitted to hospital, infected cases could either remain in the general ward until the end of their stay or go into ICU, if they became severe cases. We assumed that cases admitted in ICU entered ICU ward the same day as they were admitted in hospital (pre-ICU length of stay equal to 0 day). Once in ICU, cases could either die or stay in ICU until their discharge to general ward. Cases in general ward could either die or stay in general ward until their discharge to home.

We used age-dependent ICU admission risks for hospitalized patients and hospital and ICU death risks of hospitalized infected persons estimated by Lefrancq et al..[19] Moreover, deaths were delayed in time using the time from hospital or ICU admission to death.[19]

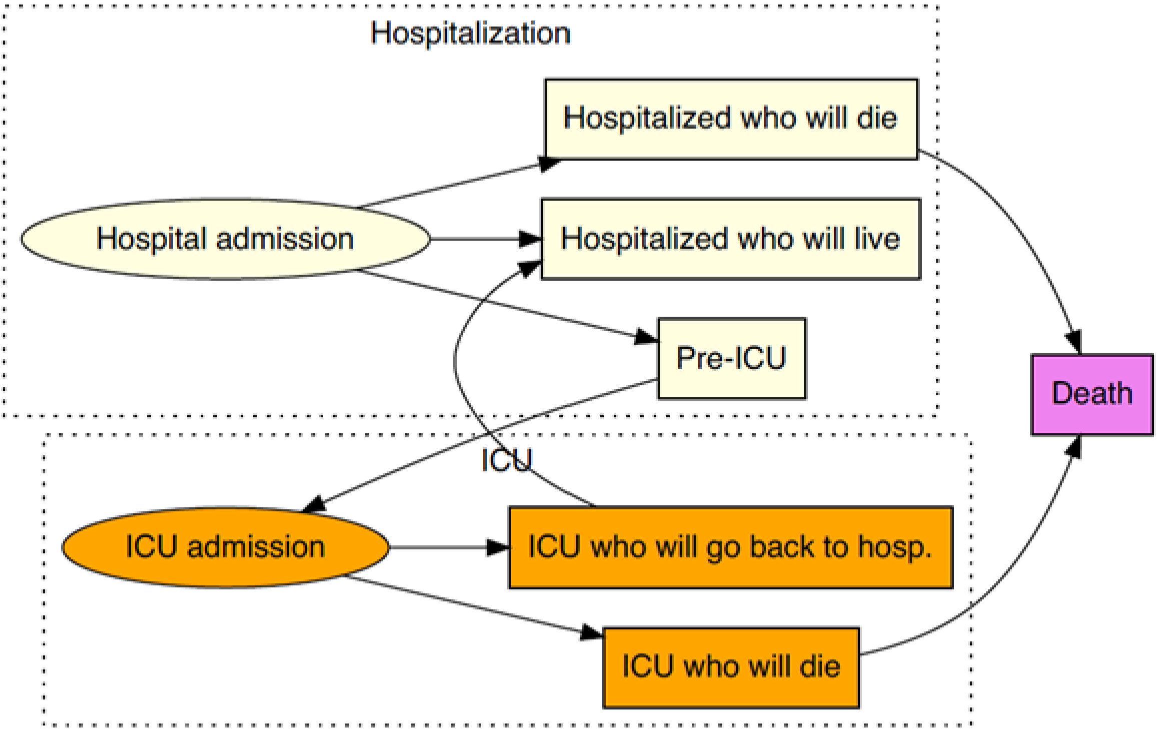


**Supplementary Figure S3** Diagram of the care pathway in hospital settings of a hospitalized COVID-19 infected individual. ICU: Intensive care unit.

### Supplementary results

#### Primary vaccination vs booster doses

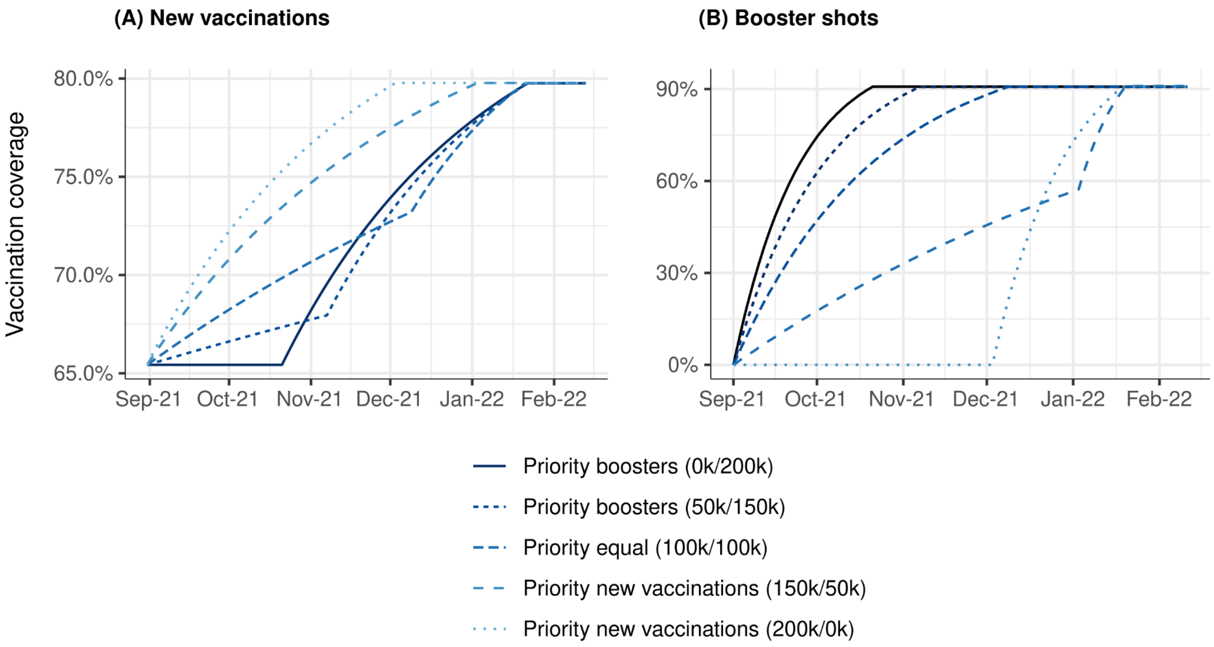


**Supplementary Figure S4** Vaccination coverage through time for each strategy from September 1st, 2021 onwards **(A)** for primary vaccinations and **(B)** for booster shots. 200,000 vaccine doses administered daily.

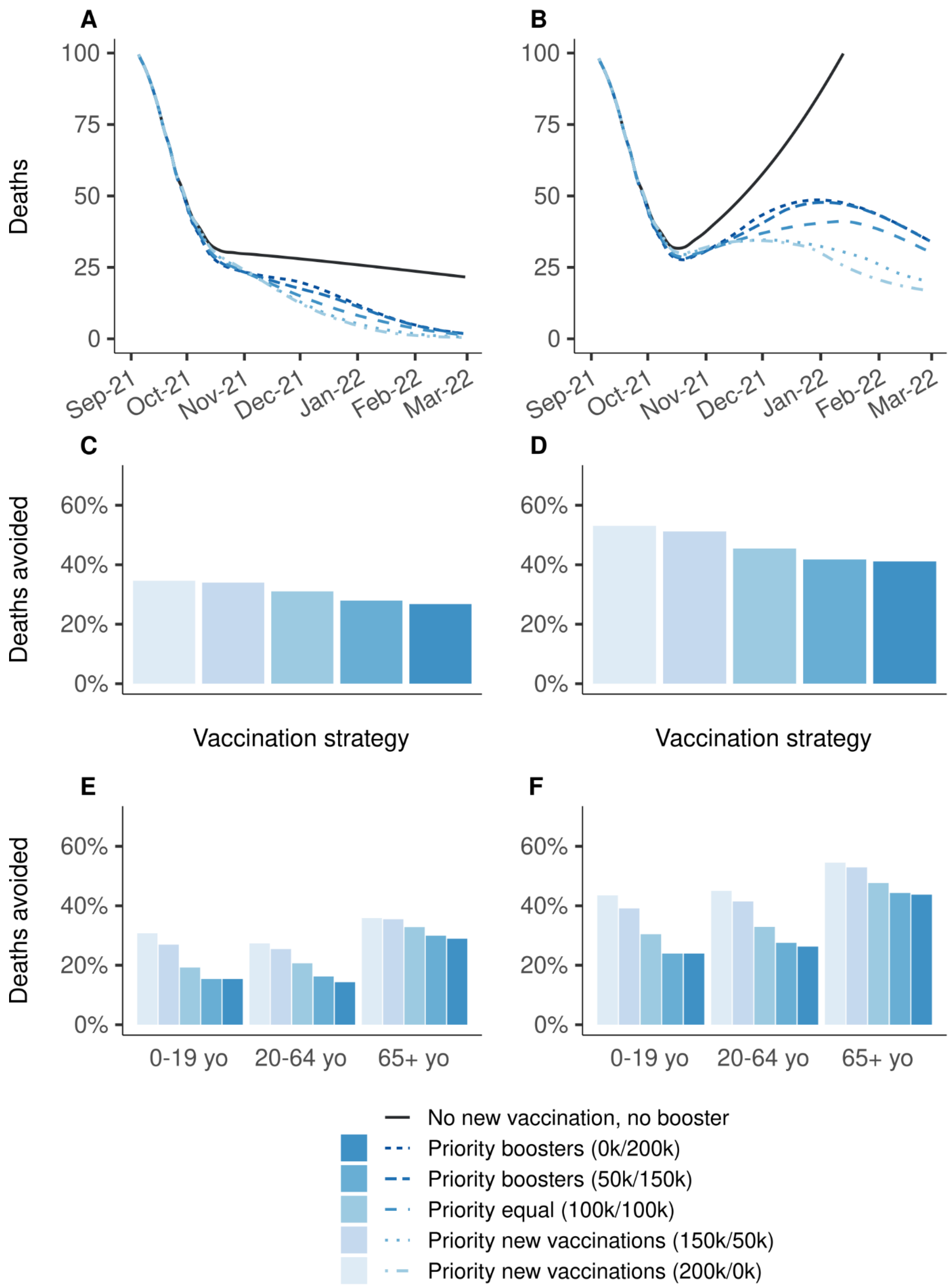


**Supplementary figure S5.** Effectiveness on the number of deaths of vaccination strategies varying in the allocation of 200,000 daily doses between primary vaccination and booster shots, over the period from September 1st, 2021 to March 1st 2022, compared to a baseline scenario in which all vaccination is stopped on September 1st, 2021. **(A, C, E)** Vaccine efficacy decreased only for people aged 65 years and older. **(B, D, F)** Vaccine efficacy decreased for all age groups. **(A,B)** Daily deaths, **(C, D)** proportion of deaths avoided, **(E, F)** proportion of deaths avoided by age group. A prioritization strategy of (150k/50k) means 150k daily doses for primary vaccination and 50k daily booster doses until 90% coverage of one target population is reached, then all 200k daily doses are allocated to the other target population.

**Sensitivity analysis with 100,000 vaccine doses administered daily**

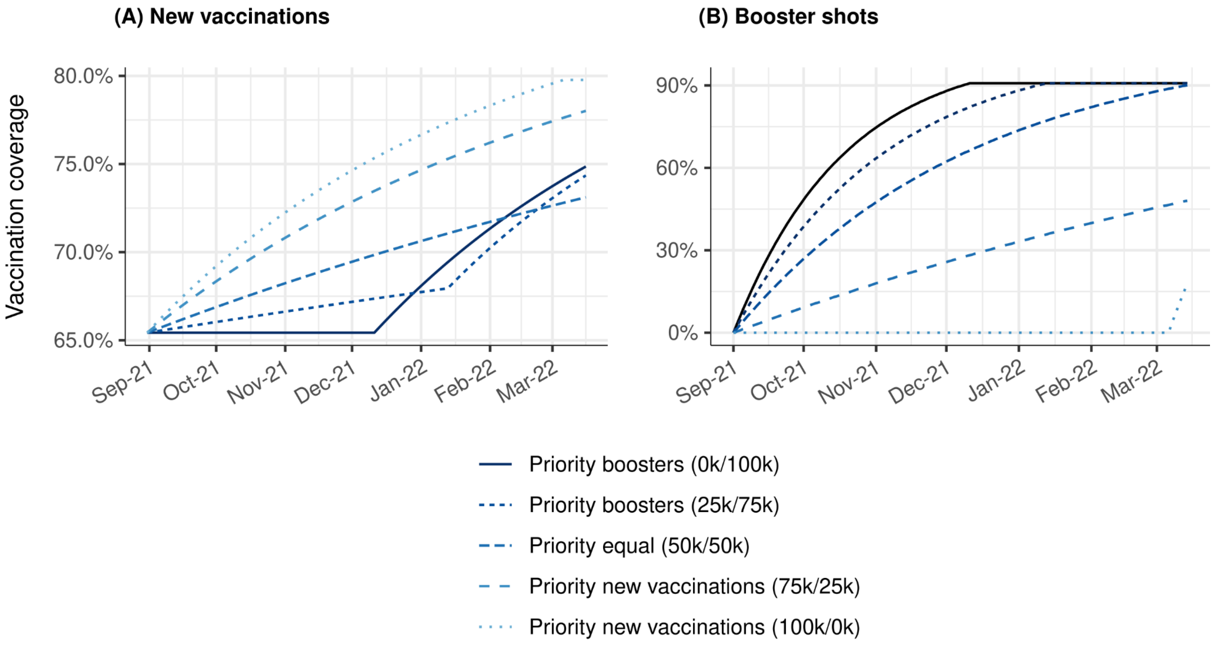


**Supplementary Figure S6** Vaccination coverage through time for each strategy from September 1st, 2021 onwards **(A)** for primary vaccinations and **(B)** for booster shots. 100,000 vaccine doses administered daily.

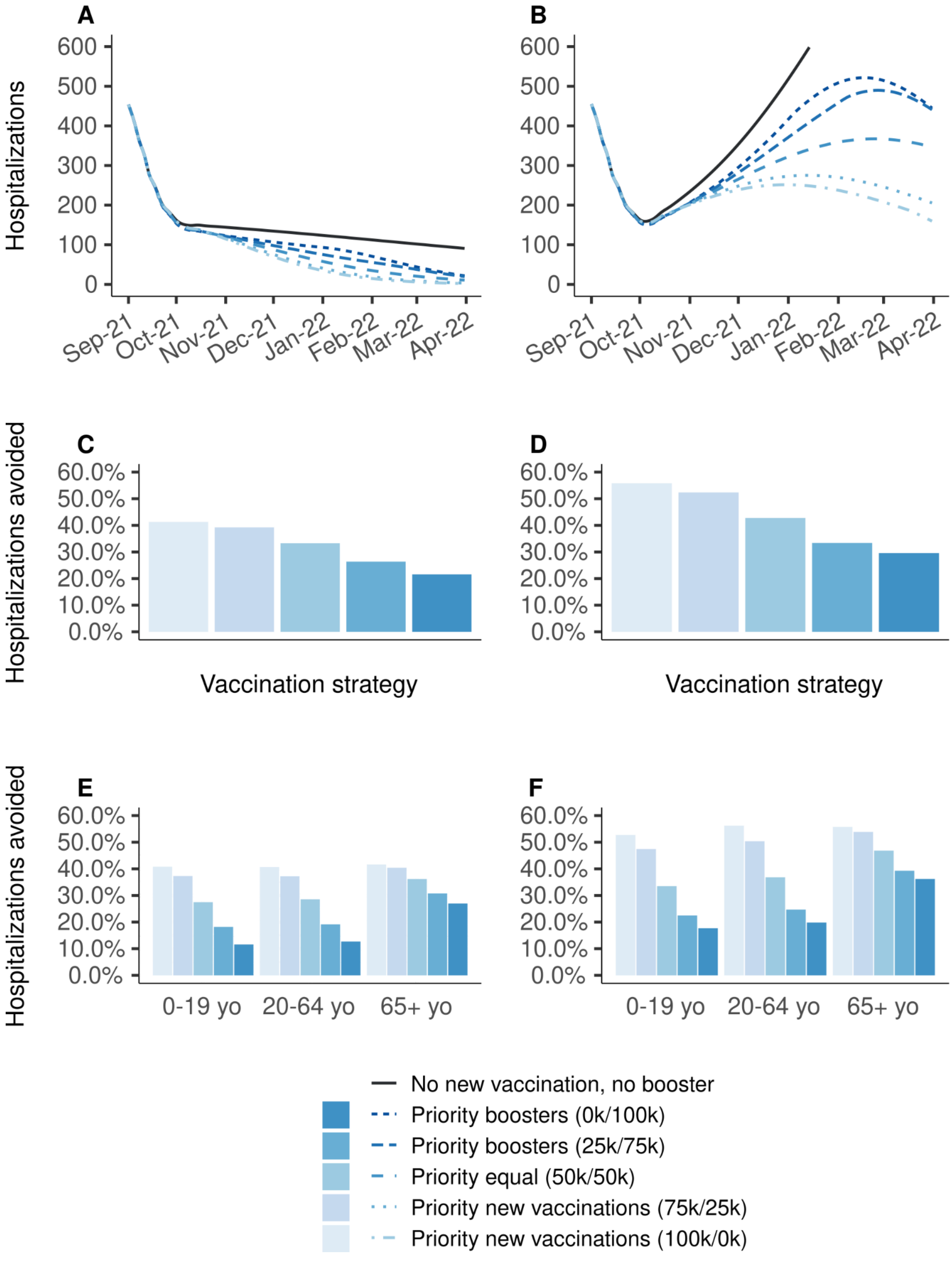


**Supplementary figure S7.** Effectiveness on hospitalizations of vaccination strategies varying in the allocation of 100,000 daily doses between primary vaccination and booster shots, over the period from September 1st, 2021 to March 1st, 2022, compared to a baseline scenario in which all vaccination is stopped on September 1st, 2021. **(A, C, E)** Vaccine efficacy decreased only for people aged 65 years and older. **(B, D, F)** Vaccine efficacy decreased for all age groups. **(A,B)** Daily new hospitalizations, **(C, D)** proportion of hospitalizations avoided, **(E, F)** proportion of hospitalizations avoided by age group. A prioritization strategy of (75k/25k) means 75k daily doses for primary vaccination and 25k daily booster doses until 90% coverage of one target population is reached, then all 100k daily doses are allocated to the other target population.

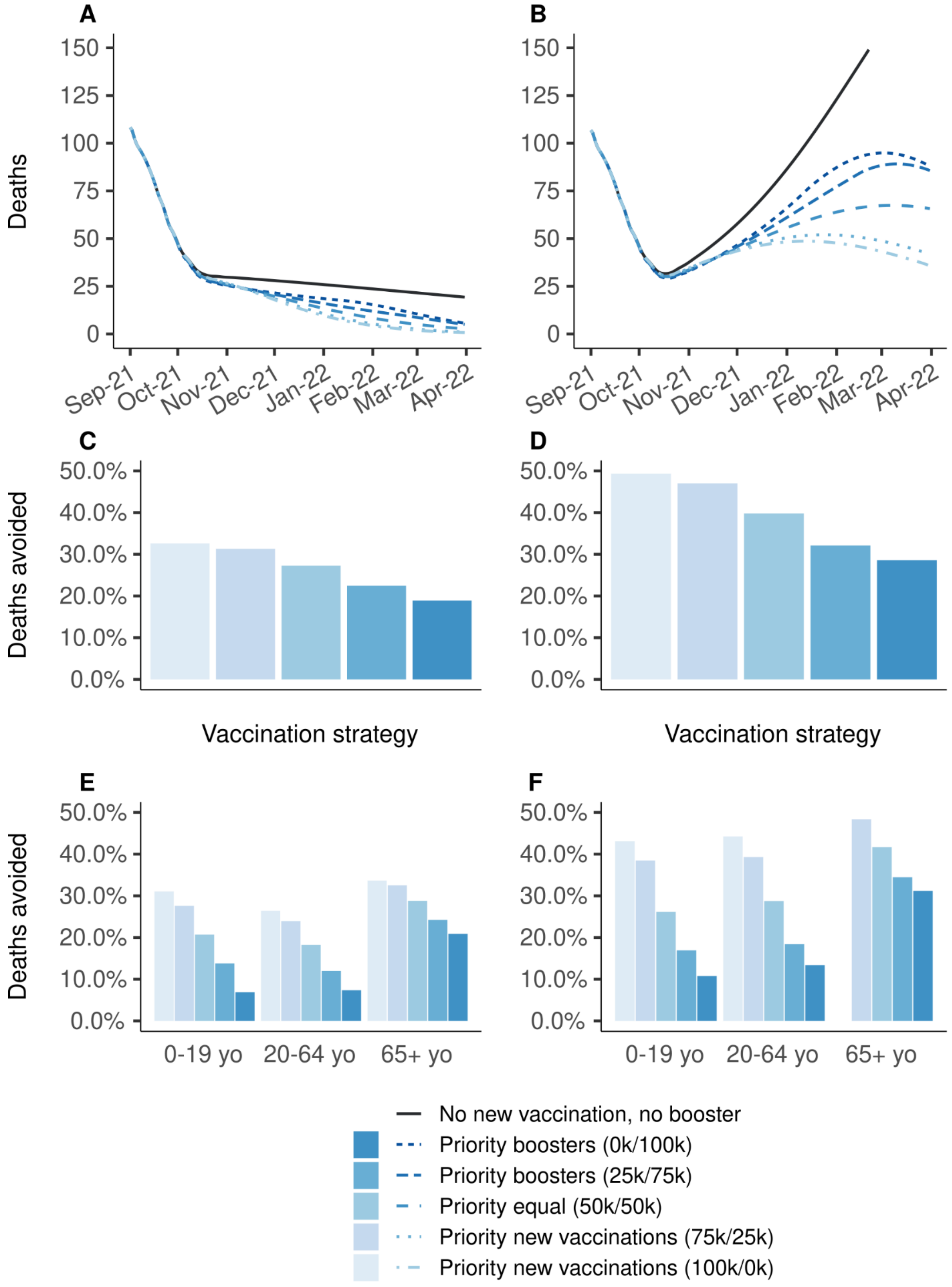


**Supplementary figure S8.** Effectiveness on the number of deaths of vaccination strategies varying in the allocation of 100,000 daily doses between primary vaccination and booster shots, over the period from September 1st, 2021 to March 1st, 2022, compared to a baseline scenario in which all vaccination is stopped on September 1st, 2021. **(A, C, E)** Vaccine efficacy decreased only for people aged 65 years and older. **(B, D, F)** Vaccine efficacy decreased for all age groups. **(A,B)** Daily deaths, **(C, D)** proportion of deaths avoided, **(E, F)** proportion of deaths avoided by age group. A prioritization strategy of (75k/25k) means 75k daily doses for primary vaccination and 25k daily booster doses until 90% coverage of one target population is reached, then all 100k daily doses are allocated to the other target population.

**Sensitivity analysis with 400,000 vaccine doses administered daily**

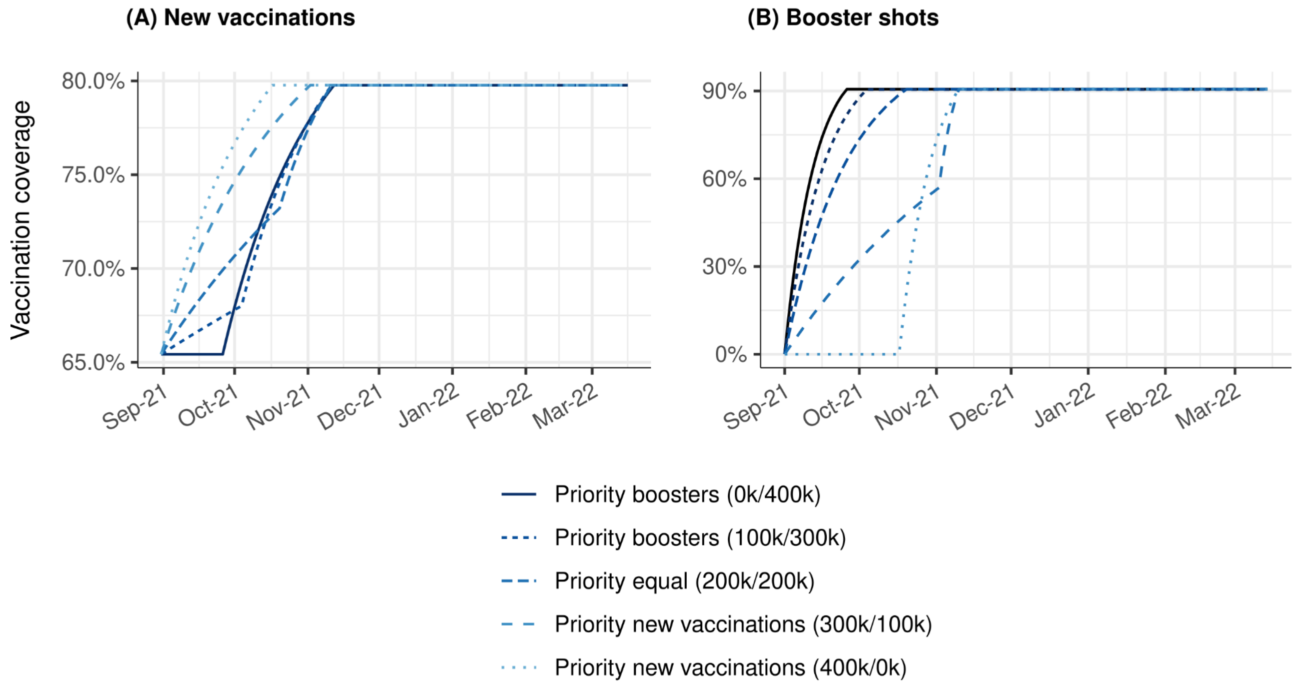


**Supplementary Figure S9** Vaccination coverage through time for each strategy from September 1st, 2021 onwards **(A)** for primary vaccinations and **(B)** for booster shots. 400,000 vaccine doses administered daily.

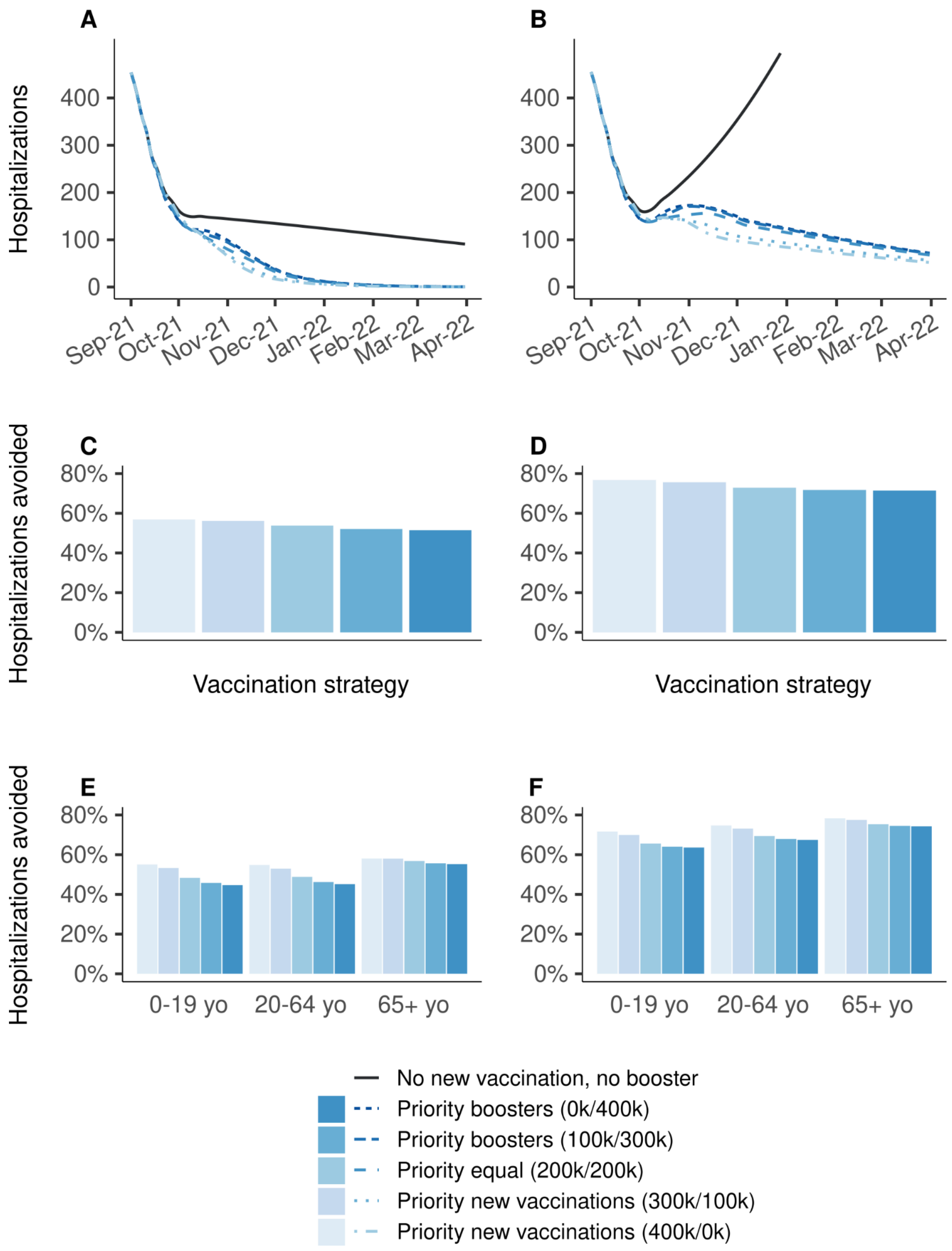


**Supplementary figure S10.** Effectiveness on hospitalizations of vaccination strategies varying in the allocation of 400,000 daily doses between primary vaccination and booster shots, over the period from September 1st, 2021 to March 1st 2022, compared to a baseline scenario in which all vaccination is stopped on September 1st, 2021. **(A, C, E)** Vaccine efficacy decreased only for people aged 65 years and older. **(B, D, F)** Vaccine efficacy decreased for all age groups. **(A,B)** Daily new hospitalizations, **(C, D)** proportion of hospitalizations avoided, **(E, F)** proportion of hospitalizations avoided by age group. A prioritization strategy of (300k/100k) means 300k daily doses for primary vaccination and 100k daily booster doses until 90% coverage of one target population is reached, then all 400k daily doses are allocated to the other target population.

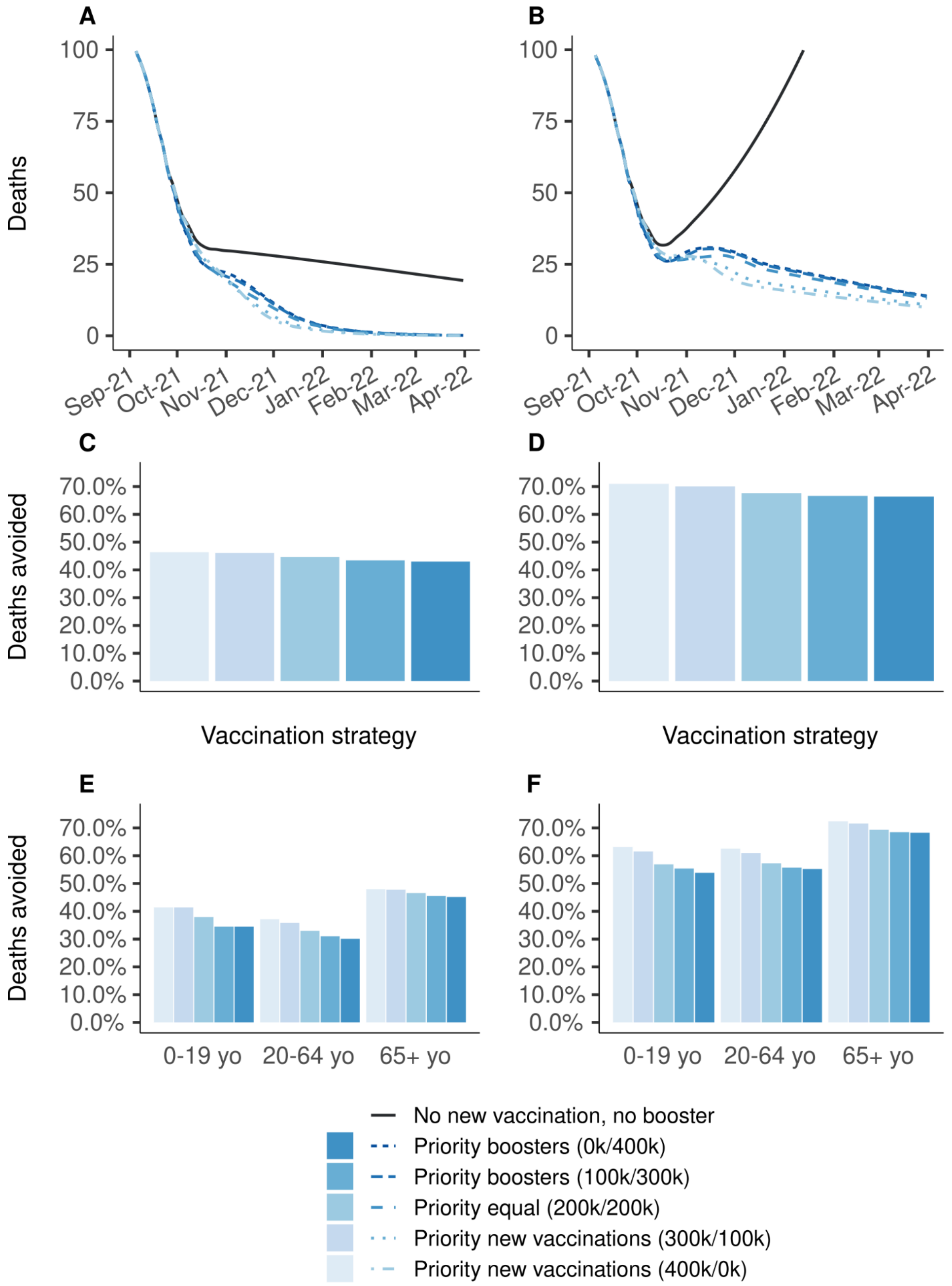


**Supplementary figure S11.** Effectiveness on the number of deaths of vaccination strategies varying in the allocation of 400,000 daily doses between primary vaccination and booster shots, over the period from September 1st, 2021 to March 1st 2022, compared to a baseline scenario in which all vaccination is stopped on September 1st, 2021. **(A, C, E)** Vaccine efficacy decreased only for people aged 65 years and older. **(B, D, F)** Vaccine efficacy decreased for all age groups. **(A,B)** Daily deaths, **(C, D)** proportion of deaths avoided, **(E, F)** proportion of deaths avoided by age group. A prioritization strategy of (300k/100k) means 300k daily doses for primary vaccination and 100k daily booster doses until 90% coverage of one target population is reached, then all 400k daily doses are allocated to the other target population.

#### Model fitting

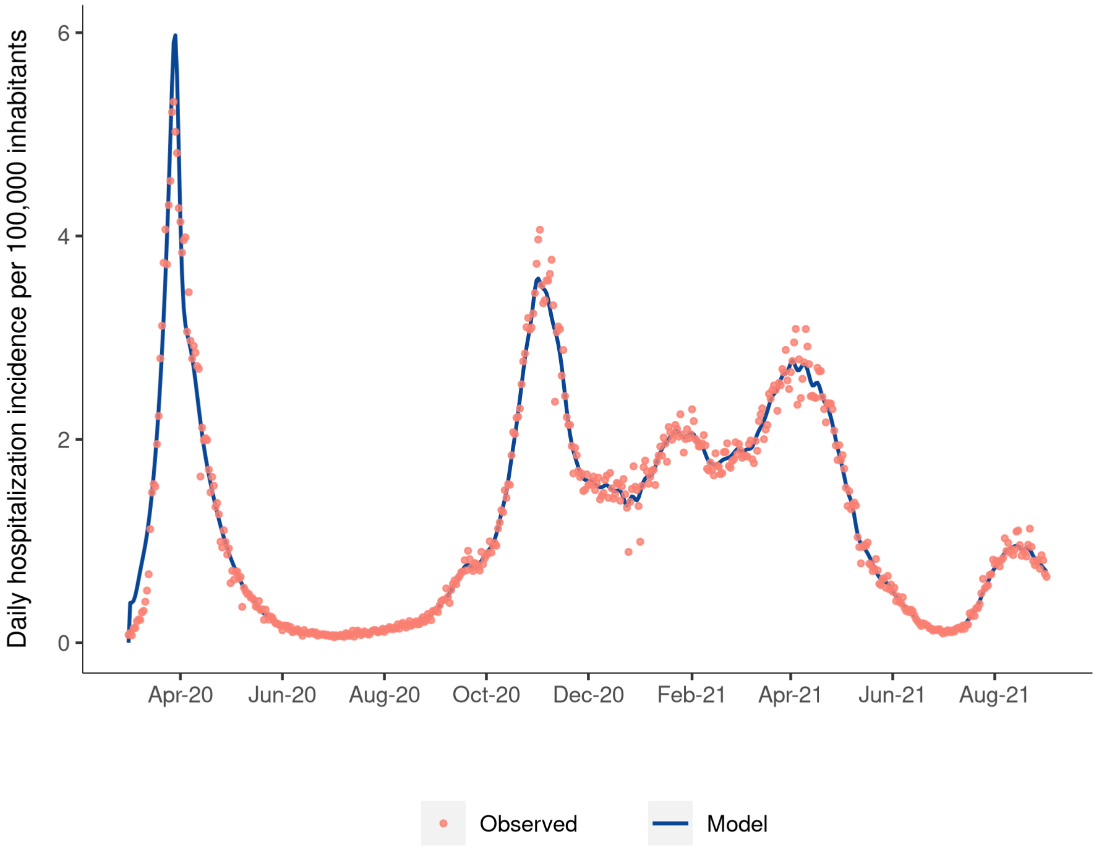


**Supplementary Figure S12** Fitting of the model to the observed daily hospitalization incidence in metropolitan France.

#### Epidemiology

Supplementary figure S13 shows the evolution of the estimated “control” and effective reproduction numbers from January 1st 2021 to August 20, 2021, for metropolitan France. The “control” reproduction number accounts for the impact of seasonality and control measures. The effective reproduction number additionally accounts for the proportion of susceptible individuals in the population. We see an increase in the reproduction numbers starting in July 2021, which is when the Delta variant of the SARS-CoV-2 virus became dominant in France. Meanwhile, the differences between the control and effective reproduction numbers increased gradually, as the proportion of susceptibles in the population decreased.

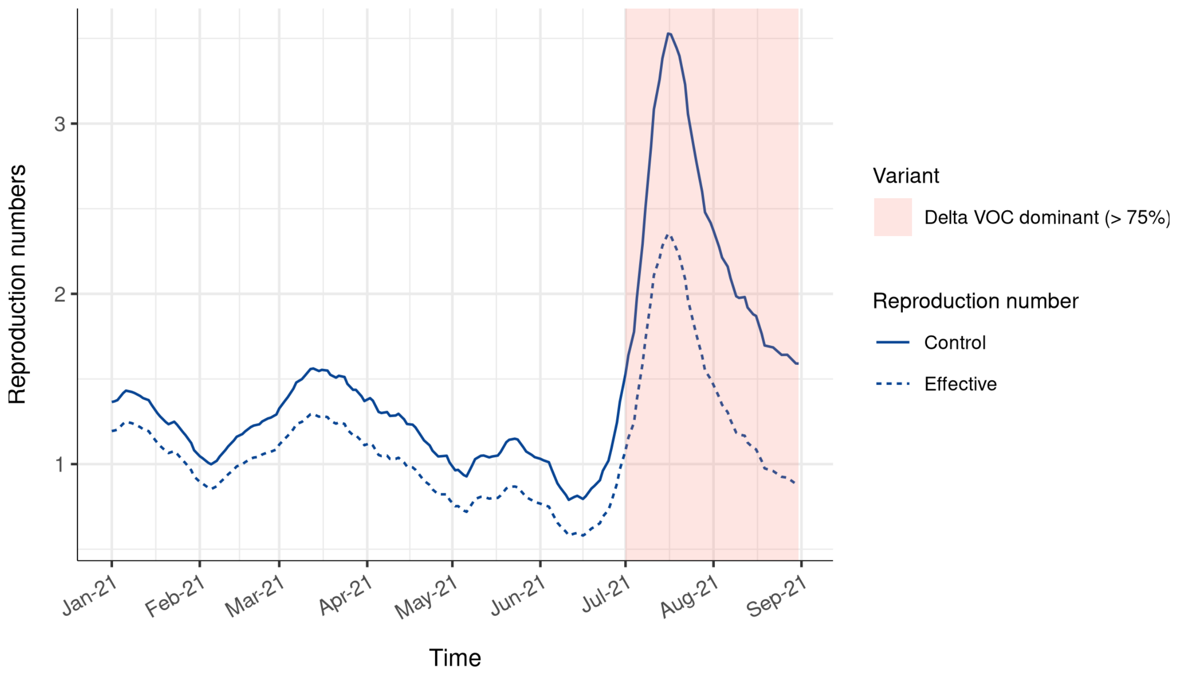


**Supplementary Figure S13.** Evolution of estimated “control” and effective reproduction numbers in metropolitan France from January to September 2021. The “control” reproduction number accounts for the impact of seasonality and control measures. The effective reproduction number additionally accounts for the proportion of susceptible individuals in the population.

Supplementary figure S14 shows the estimated proportion of the population that will have been infected by September 1st, 2021, in metropolitan France. We estimate that by that time, 20.5% of the overall population will have been infected. The most affected population are adults from 20 to 45 years old, with a seroprevalence close above 30% (31.6% for age group 20-24, 33.0% for age group 25-29, 32.3% for age group 30-34, 33.4% for age group 35-39, 31.6% for age group 40-44). This is twice as high as the estimated proportion of infected in people aged 60 years and older (16.7% for age group 60-64, 12.7% for age group 65-69, 12.5% for age group 70-74, 17.3% for age group 75-79, 9.7% for age group 80P).

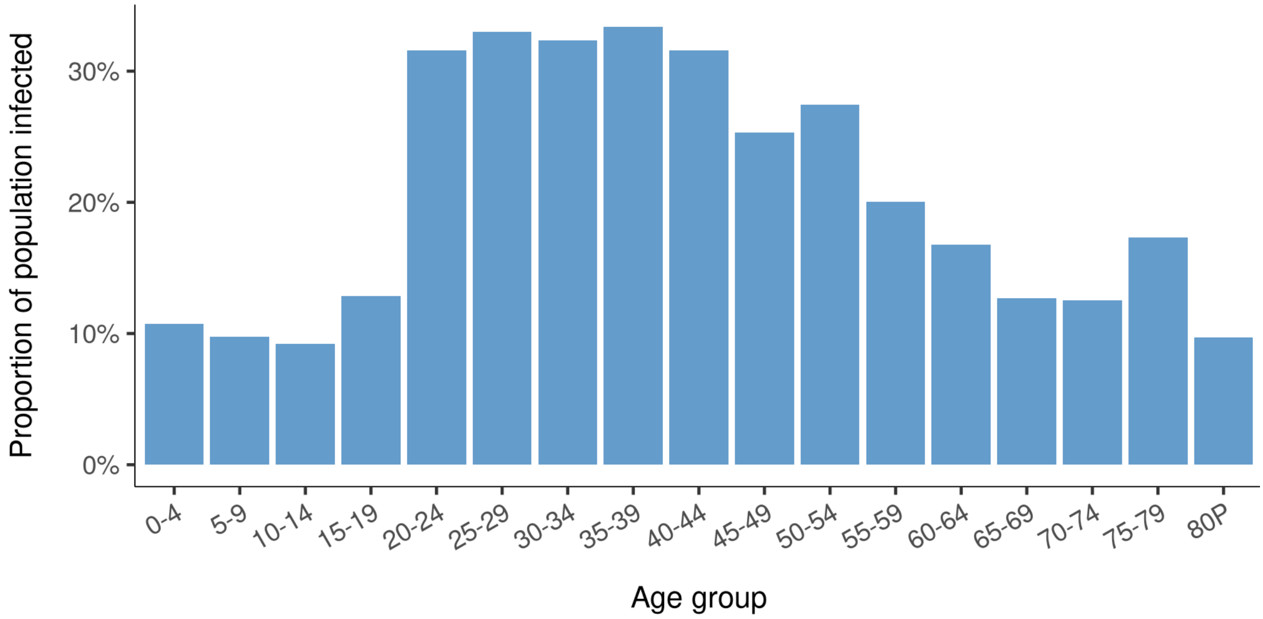


**Supplementary Figure S14**. Estimated proportion of the population that has been infected by September 1st, 2021, by age group.
